# WISE-Screen: A Smartphone-Based Analytical Framework for Automated ASD Screening and Phenotyping via High-Fidelity Eye-tracking

**DOI:** 10.64898/2026.08.21.26358650

**Authors:** Lawrence Yuk-Lung Ho, Kenneth Chi-Yin Wong, Libby Wai-Kuen Cheng, Angel Tsz-Yau Wan, Chun Hing She, Kwan-Lan Vicky Tsang, Hon-Cheong So, Stephen Kwok-Wing Tsui

## Abstract

The rising prevalence of autism spectrum disorder (ASD) strains clinical infrastructure. Gold-standard tools like ADOS-2 face high costs, specialized training requirements, and extensive waitlists, delaying diagnosis and intervention. While eye-tracking offers a promising digital biomarker, existing tools lack scalable community deployment due to hardware costs and operational constraints. Here, we introduce the WISE-Screen framework, a smartphone-based real-time architecture for autonomous ASD Screening and multidimensional phenotypic profiling, evaluating its conceptual feasibility across a development-tally diverse age range. Two machine learning pipelines processed smartphone-captured eye-gaze data: (1) a Scanpath-based (SP) pipeline utilizing saliency maps and engineered scanpath features across 34 stimuli to estimate ASD-typical gaze probabilities, and (2) a Domain-task-based (DT) pipeline evaluating responses to 17 specialized tasks across four phenotypic domains (social, emotional, sensory, executive). Models were evaluated using leave-one-out cross-validation on 35 participants (16 ASD, 19 Non-ASD, ages 2.5-17) with ADOS-2 confirmed status. Compared to a baseline demographic model (ROC-AUC = 0.82; 95% CI: 0.68-0.96), performance improved using SP model (ROC-AUC = 0.90; 95% CI: 0.78-1.00) and DT model (ROC-AUC = 0.88; 95% CI: 0.75-1.00), with the integrated model reaching a peak ROC-AUC of 0.91 (95% CI: 0.80-1.00). Age- and sex-residualized models maintained an adjusted ROC-AUC of 0.74 (95% CI:0.57-0.92), with sensory, social and emotional domains showing the strongest association. WISE-Screen offers a scalable, automated adjunct to traditional protocols, providing accessible digital phenotyping to overcome systemic ASD screening barriers, though further evaluation in larger cohorts is warranted.

## I. Introduction

AUTISM spectrum disorder (ASD) is a complex neurodevelopmental condition characterized by persistent impairment in social communication and reciprocal social interaction, alongside restricted, repetitive patterns of behaviours, interests, or activities [1, 2]. Recent epidemiological data indicates a rising prevalence; in 2020, approximately 2.76% (1 in 36) of eight-years-old children in the United States were identified with the disorder [3]. Given that early intervention is critically important for optimizing long-term developmental outcomes, research has increasingly focused on refining behavioral and physiological techniques to enhance diagnostic efficiency during early childhood [4].

The current “gold standard” for ASD diagnosis relies on resource-intensive, multidisciplinary evaluations. These protocols are centered on the Autism Diagnostic Observation Schedule, 2nd Edition (ADOS-2), a semi-structured, standardized assessment that quantifies social reciprocity and communication through direct observation of child’s interaction over a 30-60 minute duration [5]. These observations are typically supplemented by carer-proxy instruments, such as the Autism Diagnostic Interview-Revised (ADI-R) [6], to establish a comprehensive developmental history.

Despite their robust psychometric foundations, these conventional approaches face significant systemic challenges. Evaluators must undergo rigorous professional training to achieve the high inter-rater reliability, often requiring 80% agreement on protocols and algorithms, to qualify for clinical practice [7]. The complexity of these assessments renders them time-consuming, expensive and difficult to scale, leading to a persistent shortage of trained specialists and extensive waitlists. Such systemic bottlenecks often result in children missing critical developmental windows during which early intervention is most efficacious [8].

To address these barriers, researchers have increasingly explored physiological biomarkers, including genomics and electroencephalography (EEG), to augment clinical protocols [9]. Among these , eye-tracking has emerged as a promising methodology due to its capacity to quantify subtle deviations in social visual engagement [10, 11]. Distinct eye-gaze patterns are observable as early as two years of age [12]. Specifically, individuals with ASD often exhibit diminished fixation on social stimuli, such as human eyes, and a relative preference for non-social, dynamic geometric figures [12-14]. These “gaze-based” biomarkers appear to persist into adulthood, suggesting that eye-tracking provides an objective, quantifiable measure of the social-cognitive phenotypes inherent to the disorder [15].

The advantages of eye-tracking as a digital biomarker are significant. Unlike traditional clinical assessments, eye-tracking protocols are highly standardized and do not require extensive specialized training to administer. Furthermore, they yield high-dimensional, objective data that are well-suited for advanced computational analysis using machine learning (ML) techniques, such as random forest (RF), deep learning and neural networks [10, 16, 17].

Over the past decade, eye-tracking methodology has transitioned from laboratory-grade hardware to ubiquitous smartphone platforms. For instance, the iTracker has demonstrated the feasibility of using deep-learning model to predict eye-gaze positions under naturalistic conditions [18, 19]. By processing these predicted eye-gaze into automated fixations, scanpaths, and saliency maps [20-22], Startsev’s team have utilized RF models to differentiate ASD from neurotypical (non-ASD) gaze patterns [22].

While these advancements are promising, several limitations persist. Laboratory-grade hardware, such as Tobii hardware, remains prohibitively expensive, complex to set up, and lacks the portability required for broad community use [23]. Furthermore, while recent smartphone-based frameworks, developed by Chang et al. and Perochon et al, have shown potential for early ASD screening [24, 25], they often focus primarily on toddlers (ages 17 – 36 months) and emphasize binary classification (ASD vs non-ASD). Such approaches may leave older children underserved and not fully account for the phenotypic heterogeneity of the autism spectrum, which is essential for guiding personalized interventions and support. Conversely, other smartphone-based frameworks are designed to capture a broad range of behavioural features without focusing on ASD screening [9, 26].

To address these gaps, we propose the Wellmind Individual Spectrum Evaluation (WISE), a novel smartphone-based framework (WISE-Screen App) designed to integrate automated ASD screening and multidimensional phenotypic profiling. This study introduces an analytical architecture that operates across a broad developmental range (ages 2.5-17) and integrates two distinct modalities: (1) a Scanpath-based (SP) pipeline for identifying ASD-typical visual engagement, (2) a domain-task-based (DT) pipeline designed for phenotypic profiling across four behavioral domains.

Unlike previous semi-automated approach proposed by Chang et al. and Perochon et al.[24, 25], the WISE-Screen framework is designed for operational autonomy, allowing for administration by non-specialists in primary care, educational, or home settings. By emphasizing a dual-pipeline architecture, this study explores the conceptual utility of bridging the gap between ASD screening and intervention-ready phenotypic profiling by offering specialists with comprehensive data required to design personalized intervention plans. Developed through a transdisciplinary collaboration between ASD clinical experts and software engineers, this framework represents a methodological advancement toward a scalable, cost-effective adjunct to conventional instruments, potentially alleviating the systemic bottlenecks and enhancing the accessibility of ASD assessment in diverse community settings.

## II. Method

### A. Participants

Participants with ASD were recruited through open advertisements in local community newspapers, while the non-ASD control group was recruited via local community centres from students participated in extracurricular activities. Prior to participation, written informed consent was obtained from the parents or legal guardians of all minors. This study was conducted in accordance with the Declaration of Helsinki and received ethical approval from the Hong Kong Human Research independent Review Board on August 27, 2021 (Ref. No.: 2021-005). Each participant was assessed by an ADOS-certified specialist using the ADOS-2, and they were categorized into the ASD and non-ASD groups based on their ADOS-2 calibrated severity scores. To avoid data leakage during machine learning (ML) model training, ADOS-2 assessors were blinded to research outcomes.

### B. Instruments

Eye-gaze data were captured using the WISE-Screen App, deployed on an iPhone 11 (iOS 11.0 or later).(**Figure 1**). The application leverages the ARKit 2 framework to record eye-gaze coordinates and timestamps [28]. These raw data were transmitted to a secure server for preprocessing and computational analysis. Detailed hardware and software specifications for both the smartphone application and the backend server running ML analysis pipeline are provided in **Supplementary Table S1**.

**Figure 1.**
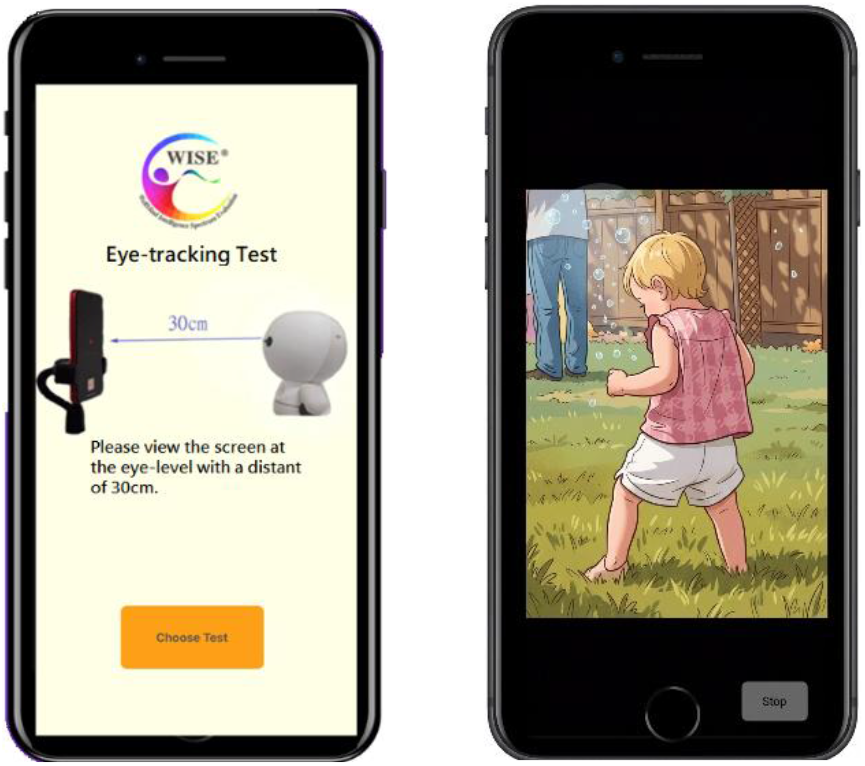
The user interface (home page) of the WISE-Screen App (Left) and a representative example of the 34 scanpath-based visual stimuli as rendered on the smartphone device (Right). The visual stimulus rendered on the smartphone (right) was extracted from the image repository curated by Duan, Zhai, Min, Che, Fang, Yang, Gutiérrez and Callet [27].

### C. Assessment Protocol

Evaluations were conducted individually in a controlled, quiet environment to minimize sensory distractions. Each participant was seated approximately 30 cm from the smartphone screen; toddlers were permitted to sit on a caregiver’s lap to ensure comfort and stability. The smartphone was secured on an adjustable mount to align with the participant’s eye level.

The assessment consisted of a series of visual stimuli and domain-specified tasks designed by a multidisciplinary team of ASD specialists, intervention experts and software engineers. The 3D face mesh technology employed in ARKit 2 obviates the need for the rigorous, active calibration procedures typically required by conventional eye-tracking hardware. To maintain eye-gaze engagement and provide a user-friendly interface, the WISE-Screen App displayed a circular reticule on the interface. While the technology was resilient to minor head movements, the assessment was repeated if significant eye-gaze data loss was detected.

### D. Scanpath-Based (SP) Pipeline for ASD Screening

#### Public Datasets and Pre-trained Models

To augment the primary study cohort, the SP pipeline incorporates two well-established external datasets for model training. The first dataset, contributed by Dorr et al. [29], provides expert-annotated eye-gaze data from 54 subjects viewing 18 high-resolution outdoor videos. This dataset was utilized to train an eye-movement profiler, facilitating the automated categorization of raw eye-gaze data into five distinct profiles: (1) fixation, (2) saccades, (3) post-saccadic oscillations, (4) smooth pursuit, and (5) noise.

The second external dataset, curated by Duan et al. [27], consists of spatiotemporal fixation sequences from 14 ASD and 14 non-ASD participants (ages 5 to 12) viewing 300 naturalistic and social images. This dataset was employed to train a RF model designed to estimate the probability of an ASD-typical eye-gaze pattern based on participant’s image-viewing behaviours. A key methodological innovation of the WISE-Screen framework is this hierarchical design: while the initial feature-extraction models were derived from external datasets (**Figure 2**, green), the final downstream SP model (**Figure 2**, purple) was evaluated exclusively within the current study cohort. By utilizing unbiased probability outputs from the external-data-derived model as inputs for the SP model, the framework minimizes the risk of model overfitting.

**Figure 2.**
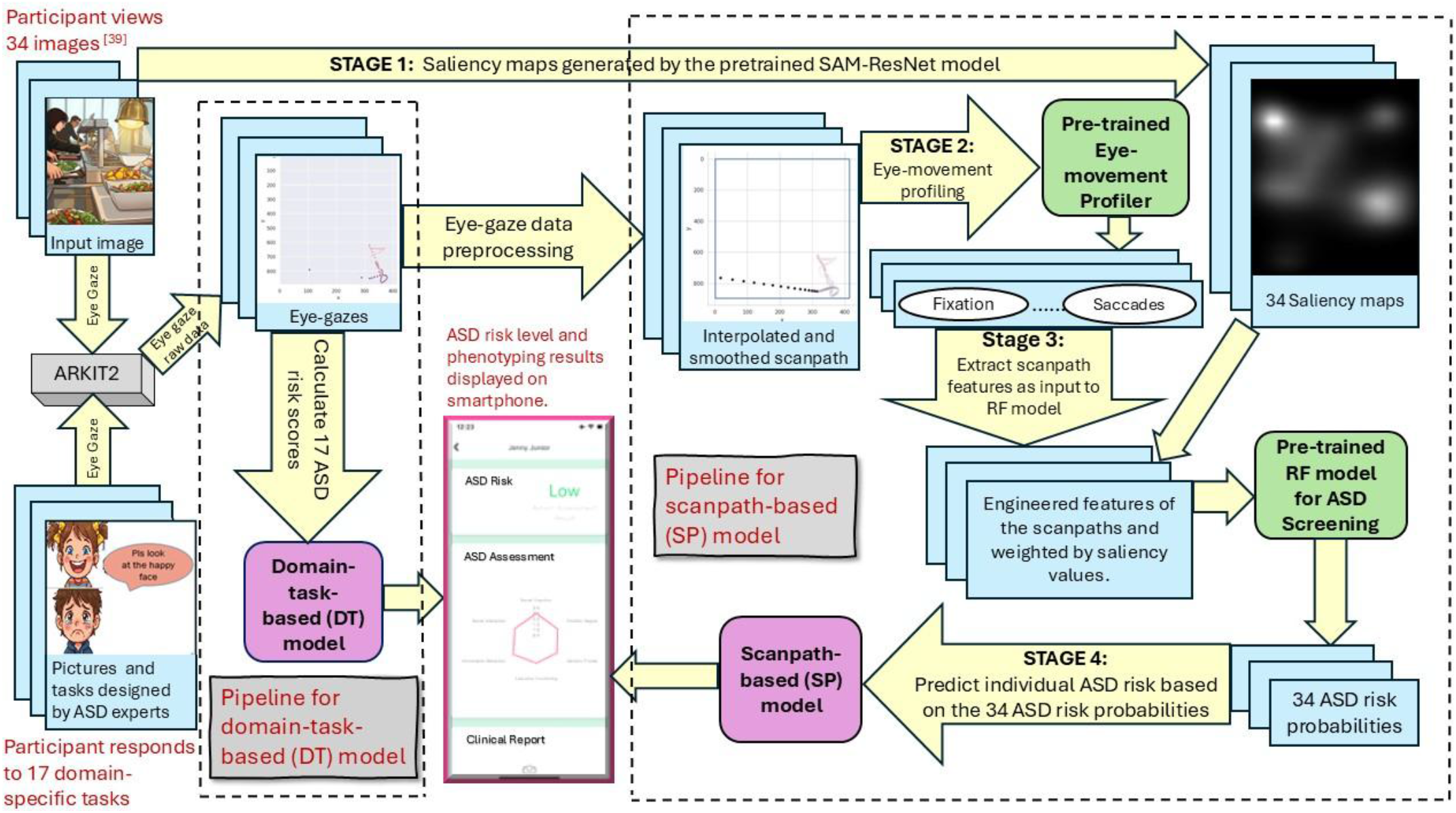
Schematic workflow of the machine learning pipelines for ASD screening and phenotypic profiling. The architecture integrates eye-gaze data from 34 scanpath-based image stimuli and 17 domain-specific tasks. Components highlighted in green represent models trained on external public datasets, while those in purple denote models trained specifically on our study cohort. Cartoon images are shown here in domain-specific tasks for illustration while real facial photographs are used in actual tasks.

Finally, the framework integrates a pretrained Saliency Attentive Model (SAM), a 50-layer Convolutional Neural Network (CNN) developed by Cornia et al. [21]. This model was used to generate high-fidelity saliency maps for the 34 visual stimuli adapted from the Duan et al. [27] dataset. These maps serve as a normative benchmark, allowing the SP pipeline to quantify captured eye-gaze deviations from the predicted saliency regions.

#### Eye-gaze Data Preprocessing

A major challenge in smartphone-based eye-tracking is the integration of heterogeneous data captured across varying sampling frequencies and hardware specifications. To address this, we design a preprocessing pipeline to address two primary technical challenges: (1) the sample rate discrepancies between data acquired via smartphone (60 Hz) and laboratory-grade trackers (up to 250 Hz) used in the external datasets [30], and (2) signal noise resulting from naturalistic head movements [29].

The framework adopts a novel distance-based approach to eye-gaze analysis, utilizing 2D Cartesian coordinate (distance between screen points) rather than conventional pixel-per-degree metrics. This approach facilitates hardware-agnostic comparisons by normalizing for variations in screen resolution and viewing distance [29]. To construct continuous trajectories from discrete eye-gaze data, we employed Cubic Hermite Spline Interpolation. For each interval [***x***_***k***,_, ***x***_***k+1***,_], the eye-gaze trajectory is modeled as a continuous function, ***p(t)*** for *t* ∈ [0, 1] (**Supplementary Figure S1**), defined as.[31]:

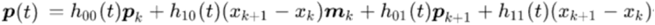

where ***p***_***k***,_ , ***p***_***k+1***_ represent Cartesian coordinates and ***m***_***k***,_ , ***m***_***k+1***_ denote their respective tangents. The interpolation is constrained by the following derivatives of the Hermite functions to ensure continuity, providing a mathematical smoothed representation of the eye-gaze path.

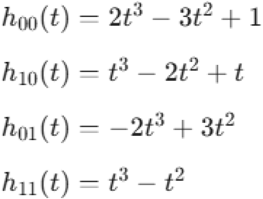

To enable integration with high-frequency public datasets, the smartphone-captured eye-gaze data were computationally upsampled from 60 Hz to 250 Hz at equidistant intervals along the function ***p(t)***. This interpolated trajectory was subsequently refined using a Savitzky-Golay filter. This specific filter was selected for its ability to minimize high-frequency artifacts introduced during the interpolation while preserving the physiological integrity of natural eye movements, such as saccadic peaks and fixation stability [32, 33]. This dual-stage approach, namely interpolation followed by localized smoothing, was implemented using scientific computing library in Python (e.g., SciPy and Scikit-Learn) [34]. Preliminary assessments of this approach (**Supplementary Figure S2**) indicate that the transition from raw 60 Hz data to a reconstructed 250 Hz signal yields a trajectory consistent with eye-movement dynamics observed in laboratory-grade equipment.

#### Constructing the Scanpath-based (SP) Pipeline

The core of the WISE-Screen framework is a novel four-stage analytical pipeline designed to translate raw eye-gaze data into an ASD risk probability. This architecture, inspired by the work of Startsev and Dorr [22], integrates deep-learning-derived visual attention maps with eye-movement profiling to characterize a typical social-visual engagement.

#### Stage 1: Saliency Maps Generation and Weighting

To quantify visual attention, the pipeline utilizes saliency maps as computational proxies for normative visual attention. These maps highlight the contextual importance of an image by emphasizing facial features and socially relevant objects (**Supplementary Figure S3A**). In this pipeline, significant deviations from these high-saliency regions are considered markers of atypical social-visual engagement, a behavior often associated with the ASD phenotypes [35].

Saliency maps for the 34 selected images were generated using a pre-trained Saliency Attentive Model (SAM-ResNet) [21] (**Supplementary Figure S3A**) .The resulting saliency distributions were converted into grayscale heatmaps (**Supplementary Figure S3B**) and subsequently transformed into a weighting matrix. This matrix assigns higher numerical weights to fixations landing on high-saliency regions (**Supplementary Figure S3B**, bright pixels) compared to those on low-saliency regions (**Supplementary Figure S3B**, dark pixels). Using the mathematical formulas detailed in **Supplementary Table S2**, this weighting approach allows for a nuanced statistical analysis of how an individual prioritizes social versus non-social information, providing a more granular metric than simple fixation counts.

#### Stage 2: Eye Movement Profiling via Expert-Rater Emulation

To achieve high-fidelity categorization of eye-gaze dynamics, we developed an automated eye-movement profiling system designed to emulate the decision-making of expert human raters. The pipeline utilizes features derived from the GazeCom dataset, a public repository of eye-movement data annotated by professional raters [29].

A RF model was trained to categorize eye-movements into five distinct profiles: fixations, saccades, post-saccadic oscillations, smooth pursuit, and noise. The model utilizes an ensemble of 1,000 trees with default hyperparameters to aggregate predictions and converge on a final profile. The model’s alignment with expert judgment was evaluated using Cohen’s Kappa coefficient to ensure inter-rater reliability and objective replicability, effectively replacing subjective manual profiling with a standardized computational process [36].

#### Stage 3: Feature Engineering and ASD Risk Probability Estimation

Following eye-movement profiling, scanpaths were constructed by connecting sequential fixation episodes. Fixations with a duration ≥ 50ms were averaged and combined, while shorter, unstable episodes or out-of-bounds fixation were excluded to ensure data integrity (**Table 1**).

**Table 1.** Illustration of the computation workflow for constructing discrete scanpaths from raw eye-gaze data. The 1^st^ table consists of raw eye-gaze data captured via the WISE-Screen App, including timestamps, Cartesian coordinates, and predicted fixation labels. In the 2^nd^ table, discrete scanpaths are constructed through the conjoining of sequential eye-gaze points. This process calculates total duration and mean spatial centroids of the scanpaths, providing the details for engineering the scanpath features required for Stage 3 of the Scanpath-based (SP) pipeline.

| Timestamp (ms) | x-coordinate | y-coordinate | Label |
| --- | --- | --- | --- |
| 340 | 254 | 467 | Fixation |
| 370 | 255 | 478 | Fixation |
| 400 | 259 | 470 | Fixation |
| 430 | 253 | 480 | Fixation |
| 460 | 248 | 475 | Fixation |
| 490 | 280 | 350 | None |
| 520 | 280 | 350 | None |
| 540 | 294 | 356 | Fixation |
| 570 | 270 | 340 | Fixation |
| 600 | 245 | 362 | Fixation |

|  | Example 1 | Example 2 |
| --- | --- | --- |
| **Duration (ms)** | $(460 - 340) = 120$ | $(600 - 540) = 60$ |
| **Mean x-coord.** | $(254 + 255 + 259 + 253 + 248) / 5 = 253.8$ | $(294 + 270 + 245) / 3 = 269.7$ |
| **Mean y-coord.** | $(467 + 478 + 470 + 480 + 475) / 5 = 474$ | $(356 + 340 + 362) / 3 = 352.7$ |

In accordance with established methodologies [22, 37], the refined scanpaths and raw eye-movement data were transformed into a high-dimensional set of engineered features (**Figure 2**). These features capture the nuances of a participant’s interaction with the visual stimuli (images), particularly regarding salient regions of interest. Critically, specific features were weighted by the heatmap matrices generated in Stage 1 to capture the degree of preference for social information — a critical differentiator between ASD and non-ASD visual behaviour. A comprehensive list of engineered features and their mathematical derivations is provided in **Supplementary Table S2**.

To develop the RF model for image-level ASD probability estimation, we utilized the Duan et al. dataset [27] (14 ASD and 14 non-ASD) across the 34 selected images. The extracted engineered features from the images, paired with diagnostic labels, were used to train an RF model (1,000 trees, a maximum depth of 10, and a minimum of 5 samples per leaf). This RF model serves as the core engine for estimating unbiased ASD-typical eye-gaze probabilities, identifying atypical eye-gaze patterns based on visual engagement with each image (**Figure 3)**

**Figure 3.**
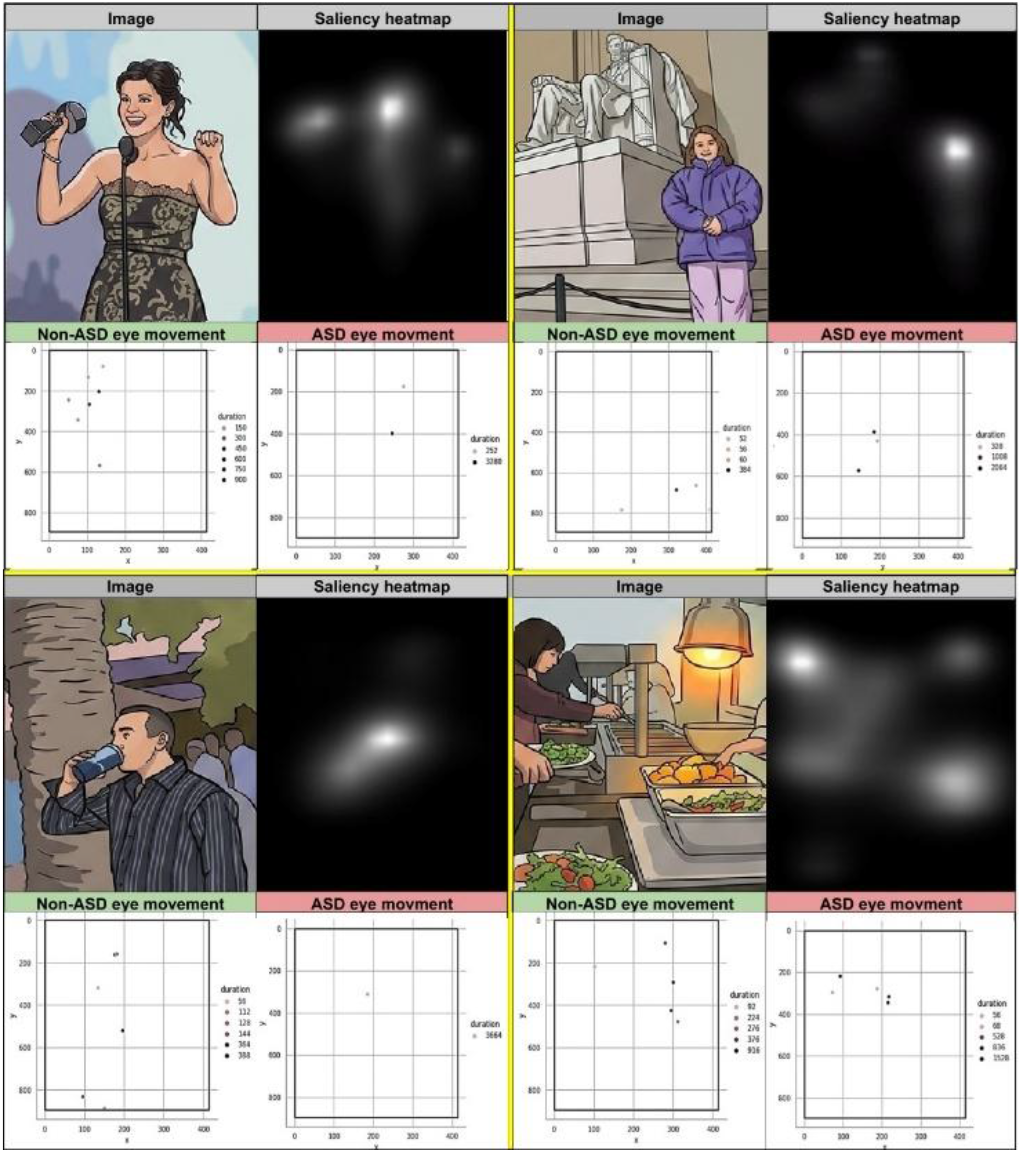
Representative examples of gaze-based fixation differences between ASD and non-ASD participants. Each of the four partitions displays a visual stimulus (upper left) alongside its computationally predicted saliency heatmap (upper right) and the fixation points for an ASD and non-ASD participant (lower plots). The figures illustrate a consistent pattern: non-ASD fixation maps show high alignment with computationally predicted social and semantic context, such as faces and central objects. Conversely, ASD fixation maps exhibit notable atypical, scattered, or peripheral eye-gaze patterns that are less aligned with these high-saliency features, illustrating the atypical visual engagement behavior in ASD. These visual stimuli were extracted from the image repository curated by Duan, Zhai, Min, Che, Fang, Yang, Gutiérrez and Callet [27].

#### Stage 4: SP model construction

The final stage of the pipeline involves applying the pre-trained RF model to our study cohort (N = 35). For each participant, a feature vector comprising 34 ASD-typical eye-gaze probabilities was generated using the pre-trained RF model derived from Stage 3. These vectors served as the input for training the downstream SP models, designed to capture higher-order relationships and complex gaze-behavioral patterns across the 34 image-viewing tasks.

A key methodological advantage of this hierarchical design is reduced risk of overfitting. Because the Stage 3 RF model was trained exclusively on an independent external dataset [27], the resulting probability vectors (and any derived summary statistics) can be treated as *out-of-sample* predictors when applied to our cohort. As a baseline, we first evaluated the predictive performance of these features by computing the arithmetic mean of the 34 ASD risk probabilities for each participant, comparing this mean against the true diagnostic labels (0 or 1) using classification metrics such as ROC-AUC.

Building upon this baseline, we developed more complex predictive models utilizing these 34 externally derived ASD risk probabilities. To optimize the final modeling architecture, we performed a systematic comparison of four ML algorithms: RF, Ridge, Elastic Net, and Lasso regression. For each algorithm, we evaluated two distinct feature sets: (1) age and sex paired with the 34 image-level ASD probabilities, and (2) age and sex paired with four derived descriptive statistics (mean, standard deviation, maximum z-score, and saturate score). The optimal architecture was selected based on cross-validated metrics ROC-AUC, with full details on hyperparameter tuning and training configurations provided in **Supplementary Table S3**.

### E. Domain-Task-Based (DT) Pipeline for Phenotypic Profiling

#### Task Design and Phenotypic Domains

The proposed DT pipeline was designed to facilitate a multidimensional phenotypic characterization of each participant. A battery of 17 domain-specific tasks was developed by our ASD specialists to capture nuances across four behavioral domains: emotional, sensory, executive, and social. This pipeline aims to identify individual phenotypic profiles, providing a conceptual basis for designing personalized intervention strategies informed by a participant’s unique distribution of strengths and challenges (**Figure 2**).

#### The Phenotypic Profiling Mechanism

The DT pipeline utilizes a preferential looking paradigm. Each task presents two distinct visual stimuli (pictures) simultaneously on the upper and lower halves of the smartphone interface. Participants are prompted to engage with either one of the pictures while the system tracks the spatial distribution of their eye-gaze in real-time.

Visual engagement is quantified by measuring the density of eye-gaze points within each of the two pictures. In this pipeline, a “phenotypic risk score” is generated based on the density of eye-gaze points directed toward the “atypical-preference” stimuli — content designed by our ASD specialists as eliciting preferential visual interest in individuals with ASD (e.g., non-social, repetitive, or geometric patterns) (**Figure 4)**. The server generates real-time risk scores for each individual task, which are then aggregated to produce a composite risk profile for each of the four phenotypic domains.

**Figure 4.**
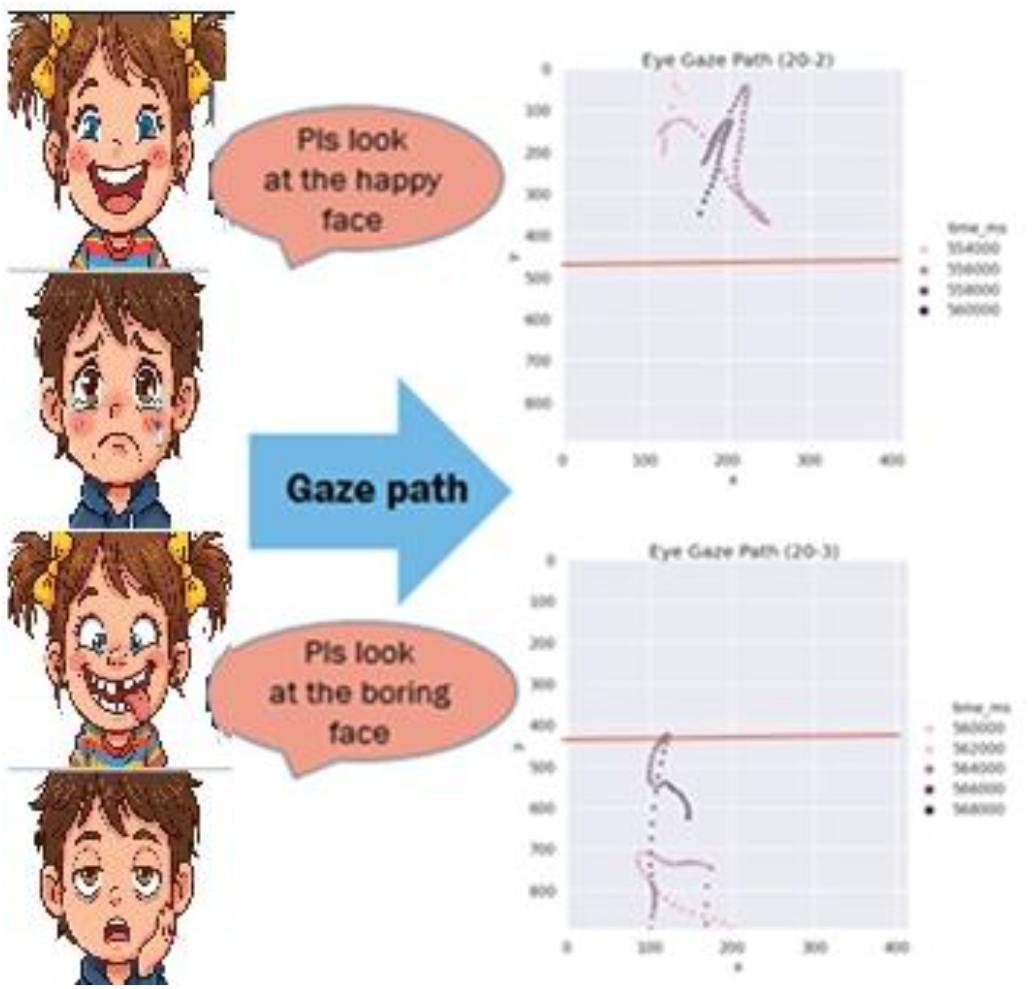
Paradigm and scoring methodology for phenotypic profiling via 17 domain tasks. The left panels illustrate two tasks in emotional domain where participants are prompted to identify target facial expressions (e.g., “happy” or “boring” faces) from pair of pictures on lower- and upper-halves of WISE- screen interface. The right panels display the corresponding temporal eye-gaze trajectories, color-coded by time (*ms*), reflecting the subject’s visual search and selection process. ASD risk score is calculated based on eye-gaze distribution; one mark is awarded if the fixation density is predominantly localized within the ASD-preferred picture, providing an objective, gaze-based measure of phenotypic response. Cartoon images are shown here for illustration while real facial photographs are used in actual tasks.

#### Evaluation of Screening Potential

As an additional analysis, we investigated the feasibility of using these phenotypic risk scores as additive features within the ASD screening workflow. We evaluated the same suite of ML algorithms employed in the SP pipeline (RF, Ridge, Elastic Net, and Lasso regression) across three distinct feature sets: (1) a combination of age, sex, and the 17 individual risk scores; (2) a domain level set incorporating age, sex, and the arithmetic means of the four phenotypic domain; and (3) a summary statistics comprising age, sex, and four derived descriptive statistics (mean, standard deviation, maximum z-score, and saturate score) derived from the 17 risk scores. This investigation aimed to determine if task-based phenotypic data provides incremental discriminative signal when integrated into the analytical framework.

### F. Statistical Analysis and Model Evaluation

To evaluate the potential of the proposed framework, predicted probabilities were compared against independently assigned clinical labels using Leave-One-Out Cross-Validation (LOOCV) approach. Given the targeted sample size (N=35), hyperparameter tuning and feature selection were strictly nested within each LOOCV fold to mitigate the risk of data leakage. In this iterative design, the model was trained on n-1 observations and evaluated on the remaining subject, ensuring that each participant served as an independent test case for the framework.

Predictive performance was quantified using several classification metrics: ROC-AUC, accuracy, sensitivity, specificity, precision, and negative predictive value (NPV). To reflect the potential utility of the framework in a real-world context, precision and NPV were adjusted to account for the estimated ASD prevalence in the general population (1:36), following the methodology proposed by Siblini et al. [38]. The optimal probability threshold for binary classification was determined via Youden’s Index [39].

We estimated 95% confidence intervals (CIs) for ROC-AUC using DeLong’s method. For other classification metrics, CIs were calculated using the Wilson score method [40], which offers greater robustness for relatively small sample sizes than the conventional Wald interval [41]. To rigorously evaluate the optimized SP, DT and combined models beyond ROC-AUC improvement (ΔAUC), we assessed added predictive power using Integrated Discrimination Improvement (IDI), and Brier score improvement. Specifically, IDI quantifies how much better the optimized model separates ASD from non-ASD cases, while Brier score improvement measures the gain in overall prediction accuracy relative to the baseline demographic model (age and sex). In these comparisons, positive values indicate that the optimized model captured more discriminative signal than the baseline demographic model. The 95% CIs and statistical significances for these comparative metrics were estimated through 1,000-iteration bootstrapping with sample replacement. All analyses were implemented using standard libraries in Python and R (**Supplementary Table S1**).

## III. Results

### A. Cohort Composition

The study cohort consisted of 35 participants (16 ASD and 19 Non-ASD), spanning a developmental range from ages 2.5 to 17 (**Supplementary Table S4**). The cohort included 20 males and 15 females. Age distribution was similar between the ASD and non-ASD groups (standardized mean difference (SMD) = 0.332, P = 0.34), whereas sex distribution differed between groups (SMD = 1.386, Fisher’s exact test P = 0.0016; **Supplementary Figure S4**). In the ASD group, 14 of 16 participants were male and 2 were female; in the non-ASD group, 6 of 19 were male and 13 were female (**Supplementary Table S4** and **Supplementary Figure S4**). ASD status was confirmed based on DSM-5 criteria [1] and validated through independent clinical assessment using the ADOS-2 [5]. To maintain a well-characterized sample for this proof-of-concept analysis, exclusion criteria included intellectual disability (IQ < 70), comorbid anxiety disorders, and ophthalmological conditions, such as visual nystagmus or uncorrected refractive errors. This pilot cohort structure provided the context for benchmarking gaze-based signatures derived from the SP and DT pipelines against demographic predictors.

### B. Demographic benchmark

To establish a reference model, we first fit a logistic model using age and sex alone. The optimized demographic model yields an ROC-AUC of 0.82 (95% CI: 0.68–0.96), an accuracy of 0.77 (95% CI: 0.59–0.89), a sensitivity of 0.68 (95% CI: 0.44–0.86), and a specificity of 0.88 (95% CI: 0.60–0.98) (**Table 2a**). Precision was 0.18 (95% CI: 0.10–0.29) and negative predictive value (NPV) was 0.99 (95% CI: 0.97–0.99). This demographic benchmark serves as the reference for evaluating whether the gaze-based signatures improved classification.

**Table 2.** Classification performance of the optimal models for ASD classification across various algorithms and feature sets. Metric cutoffs (e.g., sensitivity, specificity) were determined using Youden’s index. Performance is compared across five main configurations: **(a)** demographic baseline (age and sex) against **(b)** arithmetic mean of 34 ASD risk probabilities, **(c)** scanpath-based (SP), **(d)** domain-task-based (DT), and **(e)** integrated (Combined) models. In additional analysis. a residualized SP model was evaluated after adjusting for age and sex using leave-one-out cross-validation (LOOCV) residuals, demonstrating the intrinsic (age and sex independent) predictive validity of the gaze-based biomarkers across diverse developmental age range and imbalance sex distributions in the cohort.

| **(a) Baseline model (Ridge regression)** |  |  |  |  |  |  |
| --- | --- | --- | --- | --- | --- | --- |
| **ASD/Non-ASD ~ Age + Sex** |  |  |  |  |  |  |
|  | **ROC-AUC** | **Accuracy** | **Sensitivity** | **Specificity** | **Precision** | **NPV** |
| **Mean** | 0.82 | 0.77 | 0.68 | 0.88 | 0.18 | 0.99 |
| **95% CI** | 0.68-0.96 | 0.60-0.89 | 0.44-0.86 | 0.60-0.98 | 0.10-0.29 | 0.97-0.99 |
| **(b) 34 Scanpath-based tasks only (No age or sex)** |  |  |  |  |  |  |
| **ASD/Non-ASD ~ arithmetic mean of 34 ASD risk probabilities** |  |  |  |  |  |  |
|  | **ROC-AUC** | **Accuracy** | **Sensitivity** | **Specificity** | **Precision** | **NPV** |
| **Mean** | 0.80 | 0.77 | 0.68 | 0.88 | 0.18 | 0.99 |
| **95% CI** | 0.65-0.95 | 0.59-0.89 | 0.44-0.86 | 0.60-0.98 | 0.10-0.29 | 0.97-0.99 |
| **(c) Scanpath-based (SP) model (Ridge regression)** |  |  |  |  |  |  |
| **ASD/Non-ASD ~ Age + Sex + mean_score + sd_score + max_z_score + saturate_score** |  |  |  |  |  |  |
|  | **ROC-AUC** | **Accuracy** | **Sensitivity** | **Specificity** | **Precision** | **NPV** |
| **Mean** | 0.90 | 0.86 | 0.84 | 0.88 | 0.21 | 0.99 |
| **95% CI** | 0.78-1.00 | 0.69-0.95 | 0.60-0.96 | 0.60-0.98 | 0.13-0.33 | 0.98-1.00 |
| **(d) Domain-task-based (DT) model (Ridge regression)** |  |  |  |  |  |  |
| **ASD/Non-ASD ~ Age + Sex + mean_score + sd_score + max_z_score + saturate_score** |  |  |  |  |  |  |
|  | **ROC-AUC** | **Accuracy** | **Sensitivity** | **Specificity** | **Precision** | **NPV** |
| **Mean** | 0.88 | 0.89 | 0.89 | 0.88 | 0.22 | 1.00 |
| **95% CI** | 0.75-1.00 | 0.72-0.96 | 0.65-0.98 | 0.60-0.98 | 0.14-0.34 | 0.98-1.00 |
| **(e) Combined model (Ridge regression)** |  |  |  |  |  |  |
| **ASD/Non-ASD ~ Age + Sex + mean_SP_score + mean_DT_score** |  |  |  |  |  |  |
|  | **ROC-AUC** | **Accuracy** | **Sensitivity** | **Specificity** | **Precision** | **NPV** |
| **Mean** | 0.91 | 0.89 | 0.95 | 0.81 | 0.17 | 1.00 |
| **95% CI** | 0.80-1.00 | 0.72-0.96 | 0.72-1.00 | 0.54-0.95 | 0.11-0.26 | 0.98-1.00 |
| **(f) Residualized Scanpath-based (SP) model (Ridge regression)** |  |  |  |  |  |  |
| **ASD/Non-ASD ~ Residual(LOOCV probabilities derived from optimal SP model ~ Age + Sex)** |  |  |  |  |  |  |
|  | **ROC-AUC** | **Accuracy** | **Sensitivity** | **Specificity** | **Precision** | **NPV** |
| **Mean** | 0.74 | 0.77 | 0.84 | 0.69 | 0.14 | 0.99 |
| **95% CI** | 0.57-0.92 | 0.59-0.89 | 0.60-0.96 | 0.41-0.88 | 0.08-0.22 | 0.97-1.00 |

### C. Scanpath-based (SP) Classification

To test whether SP-derived probabilities carried discriminative information, we first summarized the 34 image-level ASD probabilities by their arithmetic means. This simple summary (without information from age or sex) reached an ROC-AUC of 0.80 (95% CI: 0.65-0.95) (**Table 2b**), which is significantly better than chance and was close to the demographic baseline ROC-AUC of 0.82. This result justified a more systematic search for a higher-performing SP model. Following a systematic evaluation of models built by four algorithms and multiple feature sets, the best-performing SP model was identified as a ridge regression with four summary statistics derived from the 34 image-level ASD probabilities (**Figure 5**). This optimized model achieved an ROC-AUC of 0.90 (95% CI: 0.78– 1.00), an accuracy of 0.86 (95% CI: 0.69- 0.95), a sensitivity of 0.84 (95% CI: 0.60–0.96), and a specificity of 0.88 (95% CI: 0.60–0.98) (**Table 2c**). Precision was 0.21 (95% CI: 0.13–0.33) and NPV was 0.99 (95% CI: 0.98-1.00). **Figure 6** indicated that, relative to the demographic baseline, the optimized SP model produced positive shifts in ΔAUC, IDI, and Brier score improvement, with confidence intervals excluding the null for these comparison metrics. These results led us to test whether DT pipeline captured a similar signal.

**Figure 5.**
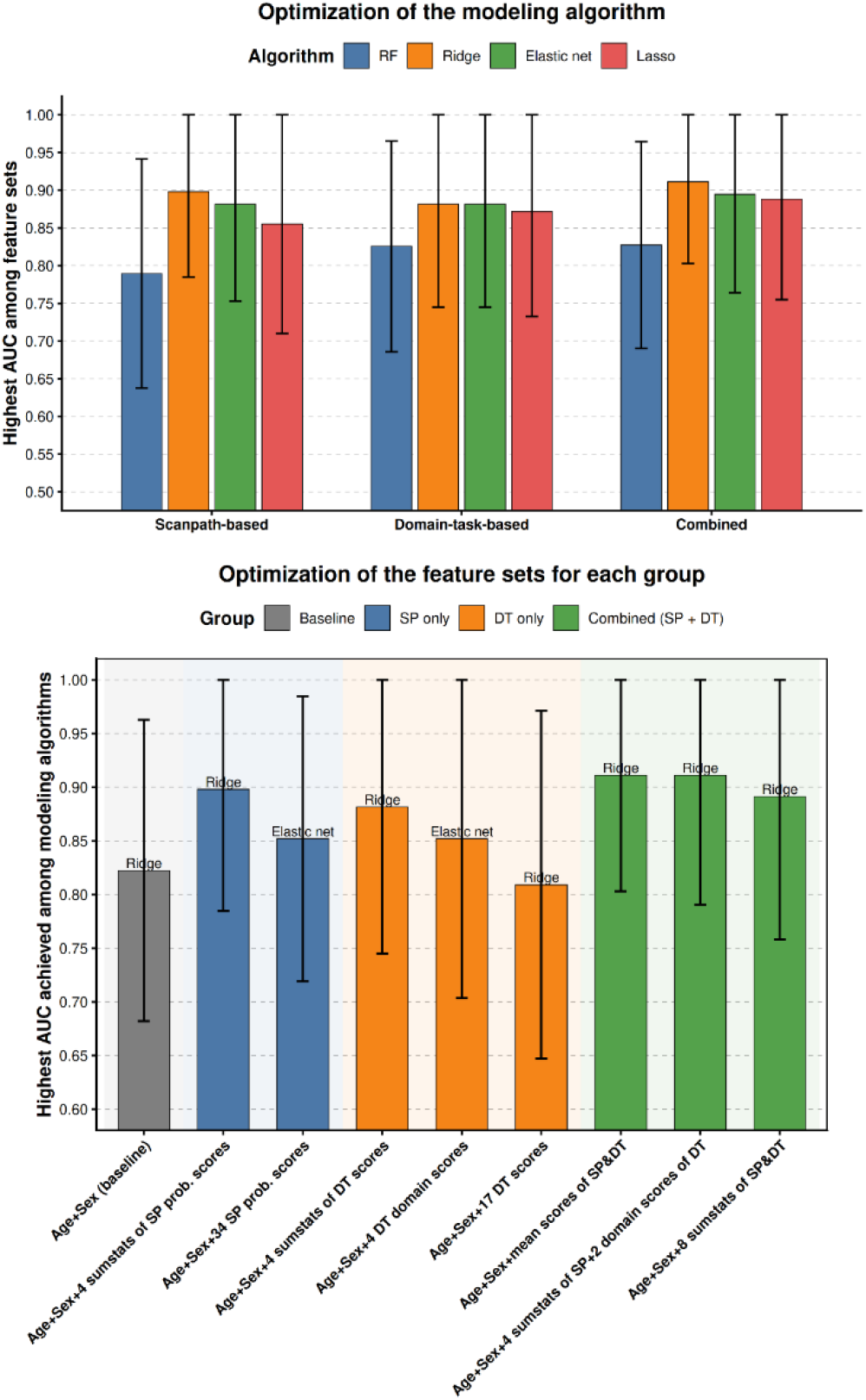
**(Top)** Comparison of modeling algorithms and feature configurations. The figure compares the predictive performance of four modeling algorithms—Random Forest (RF), Ridge, Elastic Net, and Lasso— across Scanpath-based (SP), domain-task-based (DT), and combined models. **(Bottom)** Evaluation of discrete feature sets to determine the optimal feature sets for each group. This panel compares the demographic baseline (age and sex) against varying granularities of scanpath probability and domain-task scores, with the highest-performing algorithm for each set indicated above the bar. Error bars represent 95% confidence intervals.

**Figure 6.**
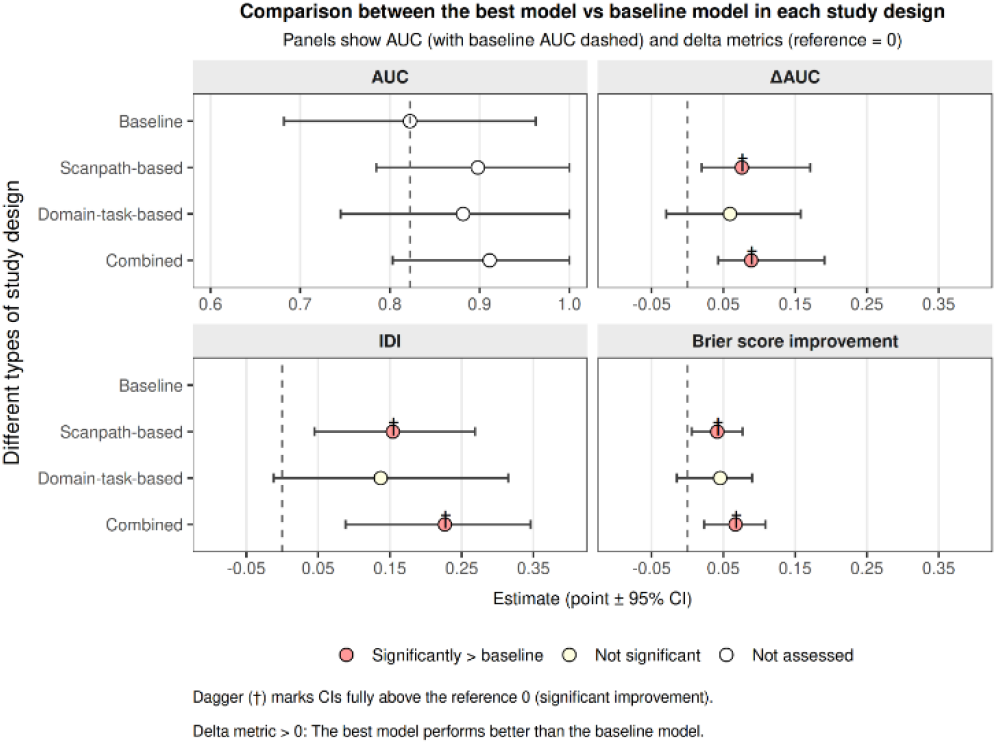
Comparative performance and incremental utility of optimized models against the demographic baseline (Age + Sex) model. The four panels display the ROC-AUC (AUC), the AUC improvement (ΔAUC), Integrated Discrimination Improvement (IDI), and Brier score improvement for the Scanpath-based, Domain-task-based, and Combined models. All performance metrics are evaluated relative to the demographic baseline (indicated by dashed vertical lines). Point estimates are presented with 95% confidence intervals (CI). Red markers and daggers () denote improvements that are statistically significant (CIs entirely above the zero-reference line), while light-colored markers () indicate non-significant differences. The results underscore the significant discriminative improvement provided by the Scanpath-based and Combined model across all comparative metrics.

#### D. Domain-task-based (DT) Classification

In this evaluation, the optimized DT model was built from four summary statistics derived from the 17 expert-designed tasks using ridge regression (**Figure 5**). This optimized DT model yielded an ROC-AUC of 0.88 (95% CI: 0.75–1.00), an accuracy of 0.89 (95% CI: 0.72–0.96), a sensitivity of 0.89 (95% CI: 0.65–0.98), and a specificity of 0.88 (95% CI: 0.60– 0.98) (**Table 2d**). Precision was 0.22 (95% CI: 0.14–0.34), and NPV was 1.00 (95% CI: 0.98-1.00). In **Figure 6**, the DT model showed point estimates above the demographic baseline, but the confidence intervals for the comparative metrics crossed the null, given the limited sample size. These results prompted us to test whether combining SP and DT information improved performance further.

### E. Integrated Model Classification

The framework’s peak observed discriminative potential was achieved through combining age, sex, with mean scores from both the SP and DT pipelines. This combined model reached the highest observed ROC-AUC of 0.91 (95% CI: 0.80–1.00), with an accuracy of 0.89 (95% CI: 0.72–0.96), a sensitivity of 0.95 (95% CI: 0.72–1.00), and a specificity of 0.81 (95% CI: 0.54–0.95) (**Table 2e**). Precision was 0.17 (95% CI: 0.11-0.26) and NPV was 1.00 (95% CI: 0.98-1.00). **Figure 6** showed that the combined model, like the optimized SP model, had positive ΔAUC, IDI, and Brier score improvement relative to the demographic baseline, with confidence intervals excluding the null. Next, we examined whether the framework also captured differences across behavioral domains.

### F. Exploratory Phenotyping Profiling

A core innovation of the WISE-Screen framework is its capacity for automated phenotypic profiling across four behavioral domains: emotional, sensory, executive, and social. Given the current lack of established benchmarks for quantifying gaze-based propensity in these specific sub- domains, we test whether it separated behavioral profiles between ASD and non-ASD groups by the ASD risk scores across the four domains. Participants with ASD had higher scores than non-ASD participants in the Social domain (P = 0.0035) and the Emotional domain (P = 0.018) (**Supplementary Figure S5**). The Sensory domain did not meet the threshold for significance (P = 0.079), and the Executive domain showed no group difference (P = 0.31) (**Supplementary Figure S5**). Next, we examined whether the model performance remained stable across age groups and after accounting for demographic covariates.

### G. Age and Sex Sensitivity

To evaluate the consistency of the optimal SP model across developmental stages, we stratified the cohort into a younger subgroup (ages 2.5–6.7 years; N = 18, 6 ASD and 12 non-ASD) and an older subgroup (ages 6.8-17 years; N = 17; 10 ASD, 7 non-ASD) (**Supplementary Table S5)**. In the younger subgroup, the demographic baseline model yielded an ROC-AUC of 0.5 (95% CI: 0.20 – 0.80), whereas the optimized SP model yielded an ROC-AUC of 0.74 (95% CI: 0.41 – 1.00). In the older subgroup, the baseline achieved an ROC-AUC of 0.80 (95% CI: 0.54 – 1.00), and the optimized SP achieved an ROC-AUC of 0.90 (95% CI: 0.73 – 1.00). In both age strata, the SP model showed higher point estimates than the demographic baseline, although the confidence intervals for comparative metrics (ΔAUC, IDI, and Brier score improvement) overlapped the null (**Supplementary Table S5)**.

In **Supplementary Figure S6**, we also evaluated the correlation between demographic factors, diagnosis, and the four SP summary statistics (mean, standard deviation, maximum z-score, and saturation score). Age showed weak correlations with the four SP summary statistics, with coefficients ranging from −0.32 to 0.32. By contrast, diagnosis correlated with sex (Spearman ρ = 0.56, ***), mean score (ρ = 0.52, **), standard deviation (ρ = 0.37, *), maximum z-score (ρ = −0.34, *), and saturation score (ρ = 0.42, *). The associations between sex and diagnosis led us to test the SP model performance after residualizing predicted probabilities for age and sex.

To investigate the potential confounding impact of the age and sex, we regressed the leave-one-out cross-validated predicted probabilities on age and sex and re-evaluated discrimination power. For the optimized SP model, the age and sex-residualized probabilities yielded an adjusted ROC-AUC of 0.74 (95% CI: 0.57-0.92) (**Table 2f**). This performance is significantly better than expected by chance, demonstrating that the eye-tracking-based model successfully predicts ASD above and beyond age and sex alone. Furthermore, as shown in **Supplementary Figure S7**, the confidence intervals for ΔAUC, IDI, and Brier score improvements between the residualized and non-residualized predictions crossed zero for the SP, DT, and combined models. This indicates that adjusting for demographic variables did not significantly degrade the models’ predictive performance.

### H. Contributions of Domain-Specific Phenotypic Scores

To delineate the contribution of each phenotypic domain scores beyond demographic predictors, we compared nested logistic regression models with Likelihood Ratio Test (LRT) against the age-and-sex baseline (**Supplementary Table S6**). The Social domain was the strongest single-domain addition (deviance = −8.0, P = 0.005). The Sensory domain also improved model (Δdeviance = −5.1, P = 0.025), as did the Emotional domain (Δdeviance = −4.3, P = 0.039), whereas the Executive domain did not (Δdeviance = −0.9, P = 0.342). The combination of Social and Sensory scores gave the largest improvement over the baseline (Δdeviance = −11.4, P = 0.003). A model that included all four domains also improved fit relative to the baseline (Δdeviance = −11.8, P = 0.019), while adding Emotional scores to a model already containing Social and Sensory scores did not improve fit further (Δdeviance = −0.2, P = 0.661). This suggests that the predictive variance captured by the Emotional domain is largely subsumed by the Social and Sensory features, making the two-domain combination the most parsimonious and effective model.

## VI. Discussion

This research explores the potential of the WISE-Screen framework as an integrated, automated architecture for the screening and phenotypic profiling of individual with ASD. Our analysis indicates that the optimized SP model achieves a notable discriminative potential, with an observed ROC-AUC of 0.90 (95% CI: 0.78–1.00). This performance suggests a substantial improvement over the demographic baseline model (ROC-AUC = 0.82; 95% CI: 0.68–0.96), as evidenced by the statistically significant improvements across all comparative metrics (**Table 2a, 2c and Figure 6**). These results illustrate the framework’s ability to extract meaning signal from high-dimensional eye-gaze data.

### A. Methodological Robustness

A distinctive strength of this framework’s design is its applicability across a broad developmental range (ages 2.5-17), a significantly wider span than the toddler-centric cohorts (age 1.5-3) assessed in previous smartphone-based eye-tracking studies [24, 25]. Our age-stratified evaluations and correlation analyses demonstrate that age does not function as a significant confounder of the SP model’s performance (**Supplementary Table S5)**. Furthermore, the four SP summary features showed weak association with age (**Supplementary Figure S6**).

Critically, a comparison of age- and sex-adjusted LOOCV-derived probabilities against non-adjusted counterparts further shows that the model’s predictive signal remains robust following residualization by age and sex (**Supplementary Figure S7**). Collectively, these findings suggest that the WISE- Screen framework may capture intrinsic, gaze-based biomarkers of social visual engagement that are conserved from early childhood through late adolescence. This stability underscores the potential of our SP pipeline to serve as a reliable screening tool across various developmental stages, bridging the gap between early detection and school-age assessment.

To reduce data leakage, the diagnostic labels were assigned independently by ADOS-2-certified assessors who were blinded to model outputs, and model development used nested leave-one-out cross-validation. In addition, the upstream RF model for estimating the image-level ASD probability was trained on an external dataset (Stage 3 of the SP pipeline) and then applied downstream to the study cohort, which further reduced the risk of overfitting to this cohort. These steps strengthen the inference that the SP model captured features linked to diagnostic status rather than measured demographic structure alone.

### B. Phenotypic Profiling and Developmental Compensation

In addition to the SP model, the Domain-Task-based (DT) pipeline demonstrated strong independent discriminative ability, achieving an impressive ROC-AUC of 0.88 and an overall accuracy of 0.89. While its incremental improvement over the demographic baseline did not reach statistical significance, likely due to the limited sample size of this pilot cohort, the DT model’s high predictive accuracy supports its utility as a potential screening mechanism. More importantly, the primary clinical value of the DT pipeline extends beyond binary classification into refined phenotypic profiling.

We hypothesize that the slight decrease in DT task performance as compared to the SP model may reflect “developmental compensation”, a phenomenon where older children with ASD develop adaptive cognitive strategies that can mask or attenuate overt atypical behaviors during structured, domain-specific tasks. In a cohort with high age-related variance, these compensatory mechanisms may narrow the observable gap in gaze behavior between ASD and non-ASD groups on certain tasks. Nonetheless, in this exploratory evaluation, the pipeline still successfully identified divergent patterns in Social and Emotional risk scores between groups (**Supplementary Figure S5**), and the inclusion of Sensory risk scores significantly enhanced the overall model fit (**Supplementary Table S6**).

The relative stability of the Executive domain scores across groups aligns with the known neurobiological heterogeneity of the autism spectrum (**Supplementary Figure S5**). Executive functioning often varies widely, ranging from significant impairment in those with co-occurring intellectual disabilities to high-level proficiency in individuals with “Asperger-type” profiles. The DT pipeline thus effectively captures the core social, emotional and sensory signatures of ASD while remaining sensitive to the diversity in executive functioning that defines the autism spectrum. By quantifying an individual’s unique strengths and challenges across these four domains, the framework introduces a novel conceptual architecture for digital phenotyping. This approach provides a potential roadmap for ASD specialists, facilitating the transition from ASD screening to the formulation of personalized, data-driven intervention strategies.

### C. Advancing the Analytical Framework: Methodological and Technological Innovations

The WISE-Screen framework introduces several methodological innovations that distinguish it from prior smartphone-based approaches [24, 25]. A primary conceptual contribution is the framework’s expansion across a broader developmental range. While existing smartphone-based screening tools have largely focused on toddlers (ages 17 to 36 months) [24, 25], the proposed analytical architecture is designed to accommodate a developmental lifespan ranging from early childhood through late adolescence (ages 2.5-17). Our evaluations suggest that the SP model may provide substantially improved discriminative potential over simple demographic variables, though the statistical magnitude of this improvement requires further investigation in larger, stratified cohorts. These initial observations suggest that the gaze-based biomarkers captured by the SP model may remain detectable across diverse age groups, maintaining a consistent signal even after adjusting for age and sex in the residualized model. This expanded developmental scope is of particular clinical interest in regions where older children often face long diagnostic waitlists.

Second, WISE-Screen is designed as a *dual-purpose* platform that integrates screening with multidimensional phenotypic profiling. Many digital tools aim primarily to produce a binary risk estimate; in contrast, WISE-Screen also provides domain-level scores (social, emotional, sensory, executive) that can support hypothesis generation and personalized intervention strategies. This linkage between screening and interpretable phenotyping is intended to narrow the practical gap between risk detection and actionable clinical insights.

From an operational perspective, the framework is engineered for a high degree of autonomy and scalability. In contrast to the semi-automated workflows of earlier systems [24, 25], the WISE-Screen architecture facilitates real-time data processing and the generation of phenotypic scores with minimal professional supervision. By streamlining the analysis pipeline, the framework offers a conceptual scalable model for deployment in diverse, non-specialized community settings (such as primary care offices and schools), potentially enhancing the accessibility of ASD services.

Technologically, the integration of the ARKit2 framework provides a robust hardware-software interface that improves upon earlier smartphone-based gaze-estimation models [19, 42]. The current framework achieves an observed uncalibrated error rate of 1.44cm (compared to 1.71cm in previous models) and a higher sampling rate (60 Hz vs. 30 Hz) [42]. These gains increase sensitivity to rapid gaze dynamics that may be informative for ASD-related attentional patterns in naturalistic smartphone use. In addition, the implementation of 3D face mesh technology of ARKit 2 and regularized Savitzky-Golay filter serves to mitigate the signal noise typically associated with naturalistic head movements during smartphone use, one of the dominant sources of error in mobile eye-tracking applications.

Beyond hardware considerations, the SP pipeline utilizes a sophisticated four-stage architecture incorporating deep-learning saliency mapping (using the SAM-ResNet model) for high-resolution visual attention analysis and saliency map generation. The framework also introduces novel data preprocessing techniques designed to harmonize eye-gaze data across disparate devices and public datasets. This methodological step is crucial, as it allows the framework to leverage diverse data sources for model refinement.

Collectively, these design choices position WISE-Screen not only as a screening tool, but as a scalable, extensible framework for gaze-based digital phenotyping in real-world settings.

### D. Limitations and Future Directions

While this research provides a promising foundation for the use of smartphone-based eye-tracking in ASD contexts, the findings should be interpreted within the constraints of a proof-of-concept work. Most notably, the limited sample size (N = 35) inherently restricts statistical power. As a result, the confidence intervals surrounding our performance metrics are relatively wide, meaning the reported estimates of model accuracy may lack precision. Consequently, these metrics should be viewed as preliminary indicators of the framework’s potential rather than definitive evidence of clinical diagnostic accuracy. In addition, the observed imbalances in sex distribution difference between the ASD and non-ASD groups and the exclusion of individuals with an IQ below 70, suggest that the current findings may not be fully generalizable to the broader autism spectrum.

Regarding the data preprocessing, while harmonizing smartphone’s eye-gaze data with high-frequency public eye-gaze datasets ensures computational consistency across disparate data sources, it remains an approximation of underlying eye-movement dynamics rather than a direct physiological reconstruction of micro-saccadic behaviours. Given this limitation, the eye-movement profiler operates on computationally harmonized trajectories rather than raw biological micro-movements. Although this does not undermine the conceptual integrity of our SP pipelines, it marks an area for future methodological refinement.

Future efforts should focus on validating this framework with larger, independent, demographically balanced cohorts, and assessing the phenotypic outputs against established clinical benchmarks to ensure the metrics are anchored in clinically meaningful constructs. Expanding the inclusion criteria to encompass individuals with wider range of cognitive profiles and common co-occurring conditions will be essential to enhance the robustness of the framework in heterogeneous, real-world clinical ASD populations. Finally, an important avenue for future research lies in the longitudinal application of these digital biomarkers. Tracking eye-gaze signals over time may offer a unique window into developmental trajectories and the efficacy of therapeutic interventions, potentially serving as an objective metric for monitoring progress. By iteratively refining this analytical architecture, the WISE-Screen framework may eventually serve as a scalable, robust, and objective adjunct to traditional clinical protocols.

## VI. Conclusion

The WISE-Screen framework represents a novel methodological advancement, integrating automated ASD screening with multidimensional phenotypic profiling via smartphone-based eye-tracking. By quantifying social-visual engagement across a broad developmental range (ages 2.5–17), this architecture offers a scalable, objective adjunct to traditional clinical protocols, potentially mitigating systemic diagnostic bottlenecks. These proof-of-concept findings are encouraging; however, they should be viewed as preliminary estimates of the framework’s potential rather than definitive clinical performance owing to the limited sample size. Future large-scale, multi-site studies are essential to establish the framework’s robustness and generalizability. Nonetheless, our work underscores the potential of high-fidelity digital screening and phenotyping to facilitate earlier ASD identification and personalized intervention strategies in diverse community settings.

## Data Availability

The participant data that support the findings of this study are available upon request from the corresponding authors, Hon-Cheong SO and Stephen Kwok-Wing TSUI. The data is not publicly available because it contains information that could compromise the privacy of research participants.

https://drive.google.com/file/d/1E6IjNmHYpP4rSouLeEFEbTyeMHcYnR4a/view?usp=drive_link

https://drive.google.com/file/d/1gg9Ir5hmdwpzhNGVKS6D7ufDNThByjGk/view?usp=drive_link

## Acknowledgment

Lawrence Yuk-Lung HO, Kenneth Chi-Yin WONG and Hon-Cheong SO designed and developed the WISE Screen App/framework. Lawrence Yuk-Lung HO, Kenneth Chi-Yin WONG and Kwan-Lan Vicky TSANG drafted the manuscript. Stephen Kwok-Wing, Hon-Cheong SO, and Kwan-Lan Vicky TSANG conceptualized, designed and supervised the study. Libby Wai-Kuen CHENG and Angel Tsz-Yau WAN recruited the participants and acquired the eye-gaze data. Angel Tsz-Yau WAN and Kwan-Lan Vicky TSANG selected the visual stimulus and designed the domain-specific tasks. Kenneth Chi-Yin WONG and Libby Wai-Kuen CHENG curated the data and conducted the statistical analyses. Chun Hing SHE designed the interface and report of the App. All authors reviewed the manuscript. Lawrence Yuk-Lung HO and Kenneth Chi-Yin WONG takes responsibility for the integrity of the data and the accuracy of the statistical analysis. This study was funded by WellMind BioMed Technology Holdings Limited. The funder played no role in study design, data collection, analysis and interpretation of data, or the writing of this manuscript. We thank all participants joining to this pilot study.

## Data Availability Statement

The participant data that support the findings of this study are available upon request from the corresponding authors, Hon-Cheong SO and Stephen Kwok-Wing TSUI. The data is not publicly available because it contains information that could compromise the privacy of research participants.

The dataset of scanpath and fixation profiles from 14 ASD and 14 non-ASD, curated by Duan, Zhai, Min, Che, Fang, Yang, Gutiérrez and Callet [27], are publicly available at http://doi.org/10.5281/zenodo.2647418. The GazeCom dataset, contributed by Dorr, Martinetz, Gegenfurtner and Barth [29], are publicly available at http://www.inb.uni-luebeck.de/tools-demos/gaze. The pre-trained SAM-ResNet model can be found in GitHub repository (https://github.com/marcellacornia/sam/).

