## Supplementary Tables for "WISE-Screen: A Smartphone-Based Analytical Framework for Automated ASD Screening and Phenotyping via High-Fidelity Eye-tracking"

**Table S1: Hardware and software specifications for the Scanpath-based (SP) and domain-task-based (DT) pipeline and smartphone application development.**

| | | | |
|---|---|---|---|
| rdware syste | Server | **CPU** | Intel Core i7 |
| | | **Total Memory** | 16GB |
| | | **Hard disk** | Total 1TB; requires 10GB free |
| | | **GPU** | GeForce RTX 2060 series |
| | App | **Smartphone** | iPhone with iOS >= v11 (equipped with "TrueDepth") |
| Software | Server (Analytical pipeline) | **Major software libraries** | TensorFlow v2.2; Keras v1.1.0; OpenCV v3.0.0; Scikit-Learn v0.23.2 |
| | | **Programming language** | Python 3.9 and R 4.5.2 |
| | App | **Tool/API** | Apple ARKit 2 |

**Table S2: Definitions and mathematical formulations of engineered features in the SP pipeline.**

| Category | Engineering features | Formula |
|---|---|---|
| General | scanpath_fix_count | N |
| | scanpath_fix_duration_ms_total | $\sum_{i=1}^{N} d_i$ |
| | scanpath_fix_duration_ms_mean | $\frac{1}{N}\sum_{i=1}^{N} d_i$ |
| Saccadic | scanpath_len_px_total | $\sum_{i=1}^{N-1}\sqrt{(x_{i+1}-x_i)^2+(y_{i+1}-y_i)^2}$ |
| | scanpath_saccade_amplitude_px_mean | $\frac{1}{N-1}\sum_{i=1}^{N-1}\sqrt{(x_{i+1}-x_i)^2+(y_{i+1}-y_i)^2}$ |
| Spatial | scanpath_distance_to_centre_px_mean | $\frac{1}{N}\sum_{i=1}^{N}\sqrt{\left(x_i-\frac{W}{2}\right)^2+\left(y_i-\frac{H}{2}\right)^2}$ |
| | scanpath_distance_to_scanpath_mean_px_mean | $\frac{1}{N}\sum_{i=1}^{N}\sqrt{\left(x_i-\frac{1}{N}\sum_{j=1}^{N}x_j\right)^2+\left(y_i-\frac{1}{N}\sum_{k=1}^{N}y_k\right)^2}$ |
| Saliency | saliency_SAM_first_fixation | $S(x_1, y_1)$ |
| | saliency_SAM_first_above_0.75*max_rank | $\min\{i \mid S(x_i, y_i) \geq 0.75 \cdot \max(S)\}$ |
| | saliency_SAM_first_above_0.9*max_rank | $\min\{i \mid S(x_i, y_i) \geq 0.9 \cdot \max(S)\}$ |
| | saliency_SAM_mean | $\frac{1}{N}\sum_{i=1}^{N} S(x_i, y_i)$ |
| | saliency_SAM_sum | $\sum_{i=1}^{N} S(x_i, y_i)$ |
| | saliency_SAM_weighted_duration_sum | $\sum_{i=1}^{N}(d_i \cdot S(x_i, y_i))$ |
| | saliency_SAM_weighted_duration_mean | $\frac{1}{N}\sum_{i=1}^{N}(d_i \cdot S(x_i, y_i))$ |
| | saliency_SAM_max | $\max_{i=1\ldots N} S(x_i, y_i)$ |
| | saliency_SAM_KLD | $D_{KL}(P\|Q)=\sum_i P(i)\log\left(\frac{P(i)}{Q(i)+\epsilon}+\epsilon\right)$ |
| | saliency_SAM_NSS | $NSS=\frac{1}{N}\sum_{i=1}^{N}\frac{S(x_i, y_i)-\mu_S}{\sigma_S}$ |

**Remarks:**

(1) $N$ - the total number of fixations in the scanpath.

(2) $d_i$ - the duration of fixation $i$.

(3) $(x_i, y_i)$ - the spatial coordinates of fixation $i$.

(4) $S$ - the saliency map, where $S(x, y)$ is the heatmap's matrix value at a specific pixel.

(5) $W, H$ - The width and height of the image viewd by participants.

(6) $KLD$ - Kullback-Leibler Divergence measures the "information loss" when using the saliency map to approximate the actual fixation map , where $P$ is the empirical fixation map and $Q$ is the predicted saliency map.

(7) NSS - Normalized Scanpath Saliency quantifies the saliency values at fixation locations after the saliency map has been standardized

(8) This table details the full suite of predictors—categorized into general, saccadic, spatial, and saliency — used in training the RF model for ASD screening in Stage 3 of the SP pipeline.

**Table S3: Final selected hyperparameters for model training across scanpath-based (SP), domain-task-based (DT) and combined models.**

| Modelling algorithm | Training parameters | |
|---|---|---|
| | Baseline model | SP/DT/Combined models |
| Random Forest | ntree=2,000, nodesize=4 | ntree=2,000, nodesize=5, mtry=4 |
| Ridge | alpha=0.0, standardize=True, type.measure="deviance" | |
| Elastic net | alpha=0.5, standardize=True, type.measure="deviance" | |
| Lasso | alpha=1.0, standardize=True, type.measure="deviance" | |

**Remarks:**

(1) ntree - Number of trees to grow

(2) nodesize - Minimum size of terminal nodes

(3) mtry - Number of variables randomly sampled as candidates at each split

(4) The best lambda is selected by minimizing the mean LOOCV error measured by model's deviance and nested within model evaluatior

**Table S4: Demographic characteristics and descriptive statistics of the ASD and non-ASD group in our cohorts.**

| Group | ASD | Non-ASD | Total |
|---|---|---|---|
| Male | 14 (70%) | 6 (30%) | 20 (57%) |
| Female | 2 (13%) | 13 (87%) | 15 (43%) |
| Total | 16 (46%) | 19 (54%) | 35 (100%) |

| Group | Age (Years) | |
|---|---|---|
| ASD | Male | Female |
| Mean | 7.6 | 10.3 |
| SD | 3.8 | 3.1 |
| N | 14 | 2 |
| Non-ASD | Male | Female |
| Mean | 7.8 | 4.6 |
| SD | 3.4 | 1.4 |
| N | 6 | 13 |

**Table S5: Age-stratified analysis of the SP and demographic baseline model performance.**

| Group | | | | | | | | | | |
|---|---|---|---|---|---|---|---|---|---|---|
| Kids (ages 2.5 - 6.7), N=18 (6 ASD and 12 non-ASD) | **Baseline model (Ridge): ASD/Non-ASD ~ Age + Sex** | | | | | | | | | |
| | **Metrics** | **ROC-AUC** | **Accuracy** | **Sensitivity** | **Specificity** | **Precision** | **NPV** | **IDI** | **Brier_diff** | **AUC_diff** |
| | **Estimate** | 0.50 | 0.67 | 0.50 | 1.00 | 1.00 | 0.97 | - | - | - |
| | **95% CI** | 0.20-0.80 | 0.41-0.86 | 0.25-0.75 | 0.52-1.00 | 0.52-1.00 | 0.93-0.99 | - | - | - |
| | **SP model (Ridge): ASD/Non-ASD ~ Age + Sex + mean_score + sd_score + max_z_score + saturate_score** | | | | | | | | | |
| | **Metrics** | **ROC-AUC** | **Accuracy** | **Sensitivity** | **Specificity** | **Precision** | **NPV** | **IDI** | **Brier_diff** | **AUC_diff** |
| | **Estimate** | 0.74 | 0.72 | 0.67 | 0.83 | 0.21 | 0.97 | 0.16 | 0.06 | 0.24 |
| | **95% CI** | 0.41-1.00 | 0.46-0.89 | 0.35-0.89 | 0.36-0.99 | 0.10-0.38 | 0.93-0.99 | -0.04-0.34 | -0.04-0.11 | -0.15-0.27 |
| Children (ages 6.8 - 17), N=17 (10 ASD and 7 non-ASD) | **Baseline model (Ridge): ASD/Non-ASD ~ Age + Sex** | | | | | | | | | |
| | **Metrics** | **ROC-AUC** | **Accuracy** | **Sensitivity** | **Specificity** | **Precision** | **NPV** | **IDI** | **Brier_diff** | **AUC_diff** |
| | **Estimate** | 0.80 | 0.88 | 1.00 | 0.80 | 0.11 | 1.00 | - | - | - |
| | **95% CI** | 0.54-1.00 | 0.62-0.98 | 0.56-1.00 | 0.44-0.96 | 0.05-0.21 | 0.98-1.00 | - | - | - |
| | **SP model (Ridge): ASD/Non-ASD ~ Age + Sex + mean_score + sd_score + max_z_score + saturate_score** | | | | | | | | | |
| | **Metrics** | **ROC-AUC** | **Accuracy** | **Sensitivity** | **Specificity** | **Precision** | **NPV** | **IDI** | **Brier_diff** | **AUC_diff** |
| | **Estimate** | 0.90 | 0.88 | 1.00 | 0.80 | 0.11 | 1.00 | 0.04 | 0.01 | 0.10 |
| | **95% CI** | 0.73-1.00 | 0.62-0.98 | 0.56-1.00 | 0.44-0.96 | 0.05-0.21 | 0.98-1.00 | -0.13-0.23 | -0.06-0.07 | -0.07-0.13 |

**Remarks:**

1. Due to small sample size, 500 iterations of bootstrapping were performed to estimate the 95% confidence intervals.

**Table S6: Likelihood ratio test (LRT) comparisons of Domain-task-based (DT) against the demographic baseline model.**

| Models with different combination of domain ASD risk scores | Baseline model | Δ Deviance | P-value (Chi-squared) |
|---|---|---|---|
| Affected ~ Sex + Age + Social + Sensory | Affected ~ Sex + Age | 11.4 | ***0.003*** |
| Affected ~ Sex + Age + Social | Affected ~ Sex + Age | 8.0 | ***0.005*** |
| Affected ~ Sex + Age + Social + Emotional | Affected ~ Sex + Age | 8.5 | ***0.014*** |
| Affected ~ Sex + Age + Emotional + Sensory + Executive + Social | Affected ~ Sex + Age | 11.8 | ***0.019*** |
| Affected ~ Sex + Age + Sensory | Affected ~ Sex + Age | 5.1 | ***0.025*** |
| Affected ~ Sex + Age + Emotional | Affected ~ Sex + Age | 4.3 | ***0.039*** |
| Affected ~ Sex + Age + Executive | Affected ~ Sex + Age | 0.9 | 0.342 |
| Affected ~ Sex + Age + Social + Sensory + Emotional | Affected ~ Sex + Age + Social + Sensory | 0.2 | 0.661 |

**Remarks**:

(1) Results are ordered by p-value to highlight the most statistically significant improvements in model likelihood.

(2) The best combination of domains is "Social + Sensory" based on these p-values.
