## Supplementary Figures for "WISE-Screen: A Smartphone-Based Analytical Framework for Automated ASD Screening and Phenotyping via High-Fidelity Eye-tracking"

#### Figure S1

Continuous trajectory interpolation using cubic Hermite polynomials. The function *p(t)* represents a third-degree polynomial constructed in Hermite form, ensuring a smooth and continuous eye-gaze path transition by interpolating between discrete data points while preserving specified endpoint positions and tangents.

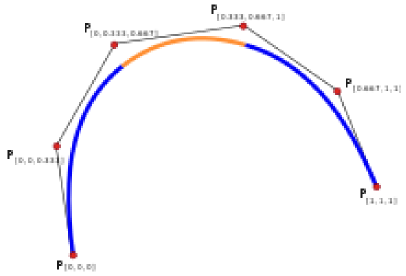


#### Figure S2

Comparison between the original 60 Hz raw data (**Left**) and the imputed 250 Hz upsampled signal (**Right**). The visualization demonstrates a reconstruction of eye-gaze dynamics, ensuring that temporal resolution and gaze trajectory characteristics remain consistent and comparable across disparate eye-tracking hardwares.

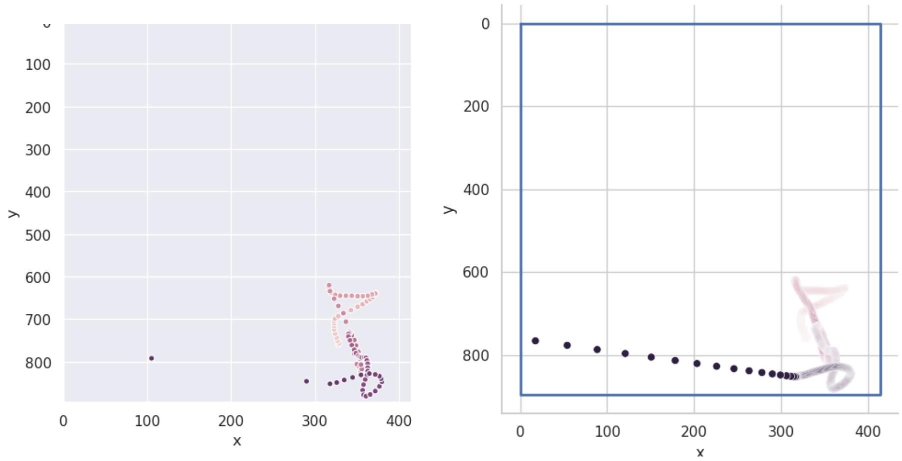

#### Figure S3

Eye-gaze data visualization and quantification using saliency maps. **(A)** Visual stimulus with an overlaid fixation heatmap, where red regions indicate peak gaze duration; the spatial distribution demonstrates the alignment between participant attention and image saliency. **(B)** Computational saliency map generated by the pre-trained SAM-ResNet model, designed to illustrate predicted spatial attention by highlighting high-contrast features and social context, such as facial regions. **(C)** Corresponding raw quantified fixation data — comprising fixation index, cartesian coordinates, and duration — providing the numerical foundation for engineering the training features of scanpath-based pipeline. These visual stimuli were extracted from the image repository curated by Duan, Zhai [29].

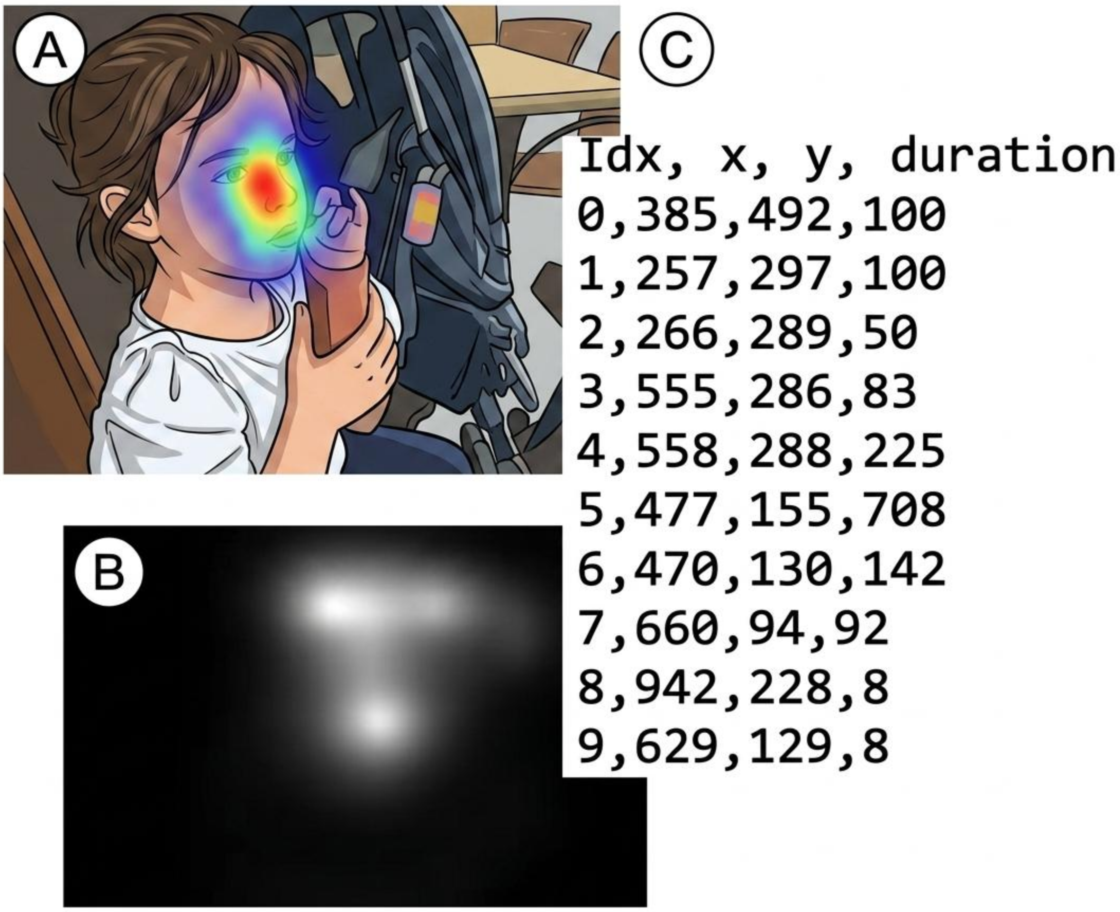

### Figure S4

The following plots show the age and sex discrepancy between Non-ASD vs ASD group

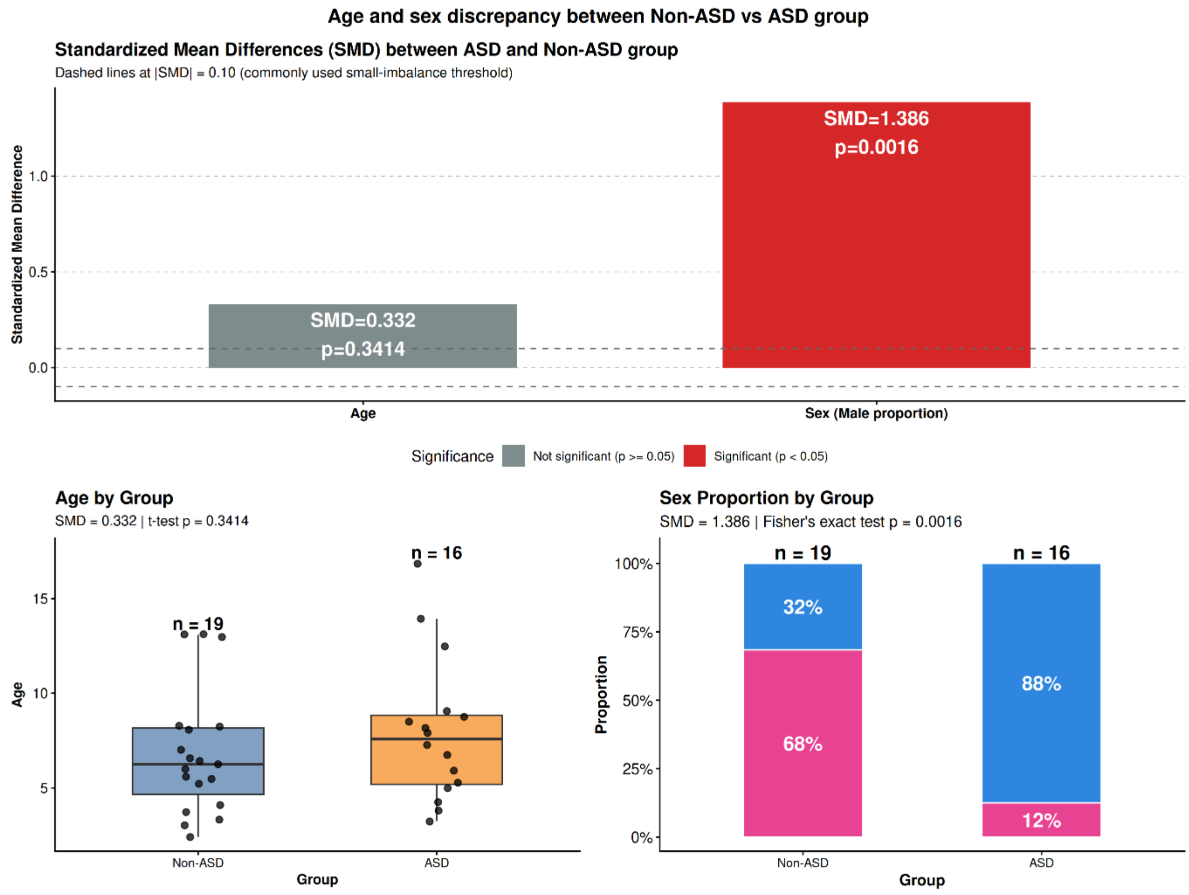

#### Figure S5

Group differences in ASD risk scores across four domains. Results of one-tailed independent samples t-tests comparing ASD and non-ASD groups across four domains. ASD participants exhibited significantly higher risk scores ($p < 0.05$) compared to the non-ASD group in the Emotional and Social domains, whereas no significant differences were observed in the remaining domains. Error bars represent standard errors of the mean.

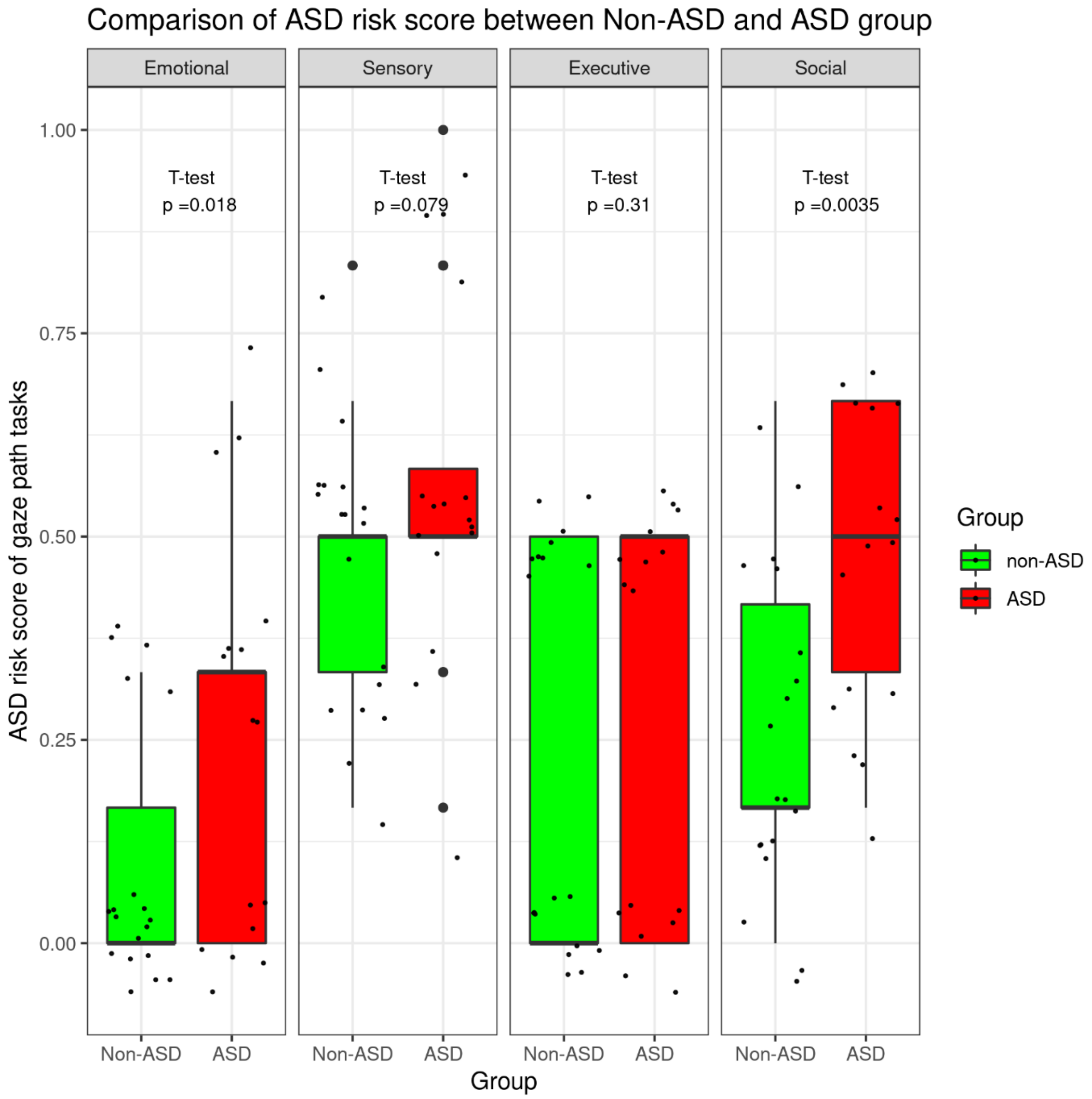

### Figure S6

Spearman correlation matrix of ASD diagnosis, demographic variables, and scanpath-based (SP) features. This heatmap illustrates the associations between ASD diagnosis, demographic factors, and the four summary statistics utilized in the optimized SP model (Mean, SD, Max Z-score, and Saturate score). Notably, age showed weak, non-significant correlations with the SP summary features, suggesting limited age-related confounding within this cohort. Conversely, the four summary statistics demonstrate significant correlations with diagnostic status ($P < 0.05$), underscoring their discriminative capability in ASD screening.

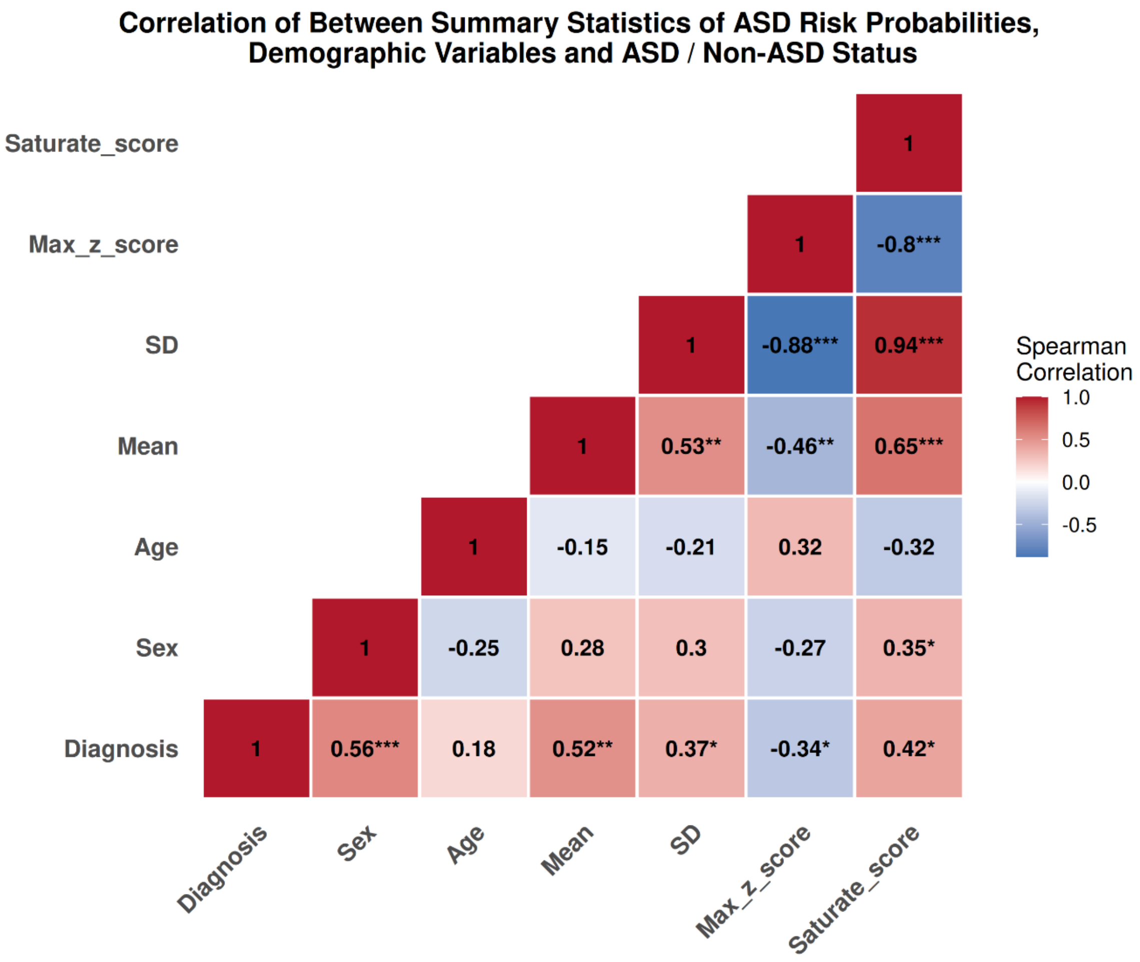


Spearman correlation coefficients are shown. Statistical significance is indicated as:
* p < 0.05; ** p < 0.01; *** p < 0.001.

#### Figure S7

Impact of age and sex residualization on model predictive performance. Comparative analysis of classification accuracy for models with and without demographic (age and sex) residualization by regressing the leave-one-out cross-validated predicted probabilities on age and sex and re-evaluated discrimination power. The plots illustrate the extent to which model performance is preserved after regressing out variance associated with age and sex, thereby showing the intrinsic predictive power of the gaze-based biomarkers.

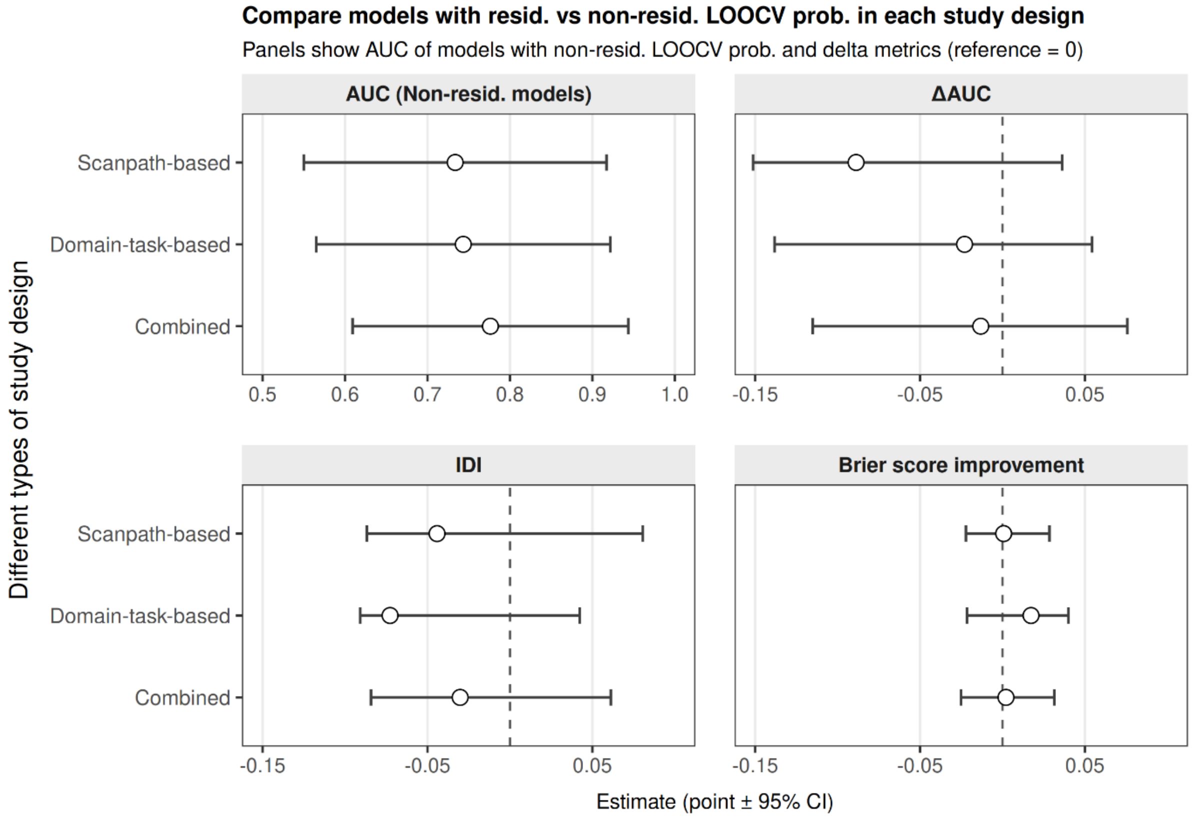


Overlapping 95% CI with the reference line indicate the predictive performance does not differ significantly between features residualized by Age & Sex and those left unadjusted.
